# A phase I clinical trial of intrahepatic artery delivery of TG6002 in combination with oral 5-fluorocytosine in patients with liver-dominant metastatic colorectal cancer

**DOI:** 10.1101/2024.07.30.24311046

**Authors:** Emma J. West, Alain Sadoun, Kaidre Bendjama, Philippe Erbs, Cristina Smolenschi, Philippe A. Cassier, Thierry de Baere, Sophie Sainte-Croix, Maud Brandely, Alan A. Melcher, Fay Ismail, Karen J. Scott, Angela Bennett, Emma Banks, Ewa Gasior, Sarah Kent, Marta Kurzawa, Christopher Hammond, Jai V. Patel, Fiona J. Collinson, Chris Twelves, D. Alan Anthoney, Dan Swinson, Adel Samson

## Abstract

**Background:** Effective treatment for patients with metastatic cancer is limited, particularly for colorectal cancer patients with metastatic liver lesions (mCRC), where accessibility to numerous tumours is essential for favourable clinical outcomes. Oncolytic viruses (OVs) selectively replicate in cancer cells; however, direct targeting of inaccessible lesions is limited when using conventional intravenous (*i.v.*) or intratumoural (*i.t.*) administration routes.

**Methods:** We conducted a multi-centre, dose-escalation, phase I study of vaccinia virus, TG6002, via intrahepatic artery (IHA) delivery in combination with the oral pro-drug 5-fluorocytosine (5-FC) to fifteen mCRC patients.

**Results:** Successful IHA delivery of replication-competent TG6002 was achieved, as demonstrated by virus within tumour biopsies. Functional transcription of the *FCU1* transgene indicates viral replication within the tumour, with higher plasma concentrations of 5-fluorouracil (5-FU) associated with patients receiving the highest dose of TG6002. IHA delivery of TG6002 correlated with a robust systemic peripheral immune response to virus with activation of peripheral blood mononuclear cells, associated with a proinflammatory cytokine response and release of calreticulin, potentially indicating immunogenic cell death. Gene Ontology analyses of differentially-expressed genes reveal a significant immune response at the transcriptional level in response to treatment. Moreover, an increase in the number and frequency of T cell receptor clones against both cancer- and neo-antigens, with elevated functional activity, may be associated with improved anti-cancer activity. Despite these findings, no clinical efficacy was observed.

**Conclusions:** In summary, these data demonstrate delivery of OV to tumour via IHA administration, associated with viral replication and significant peripheral immune activation. Collectively, the data supports the need for future studies using IHA administration of OVs.

**SUMMARY:** A phase Ia study of TG6002 oncolytic vaccinia virus administration via the hepatic artery in patients with colorectal cancer liver metastases. Virus was delivered to tumour with functional activity of the virus-encoded *FCU1* transgene, eliciting innate and adaptive anti-cancer immunity.

## BACKGROUND

Colorectal cancer (CRC) is a leading cause of cancer-associated deaths in Western populations and the third most frequent cause of cancer-related deaths worldwide [1]. The five-year survival rate for localised disease is approximately 91 %; however, around 25-30 % of patients with CRC develop liver metastases [2], which is associated with a 5-year survival rate of only 13 % [3]. For these patients, systemic anti-cancer therapy is the mainstay of treatment, with 5-fluorouracil (5-FU) being commonly employed either as monotherapy or in combination with other cytotoxics. However, 5-FU has limitations including intravenous (*i.v.*) administration, short half-life, significant systemic toxicity and drug resistance [4]. For patients with liver-dominant metastatic CRC (mCRC), locoregional therapies offer the prospect of effective treatment, whilst limiting systemic toxicity.

Oncolytic viruses (OVs) are principally immunotherapeutic agents that preferentially replicate in malignant cells, ultimately inducing immunogenic cell death (ICD). OVs can be engineered to express transgenes with immune-stimulating functions or highly specific downstream targets [5]. Many engineered OVs have been evaluated in randomised trials, with three currently licensed as standard care [6]. One virus that has been tested extensively in the clinical setting is *Pexa-Vec* (*Pexastimogene Devacirepvec*; JX-594, TG6006), an engineered Wyeth-strain vaccinia virus [7] developed by Transgene and Sillagen. Clinical efficacy as a single agent by intratumoural (*i.t.*) injection was observed in a dose-comparison, randomised study in patients with hepatocellular carcinoma (HCC), where overall survival was significantly longer for patients in the high dose group [8]. Furthermore, *i.v.* delivery to tumour is also possible at a dose of 1×10^9^ plaque-forming units (pfu) [9].

TG6002 was developed by engineering the highly oncolytic Copenhagen vaccinia strain [10,11], incorporating gene modifications to enhance its anti-tumour activity and clinical impact.

Thymidine kinase (*TK*) and ribonucleotide reductase (*RR*) genes are deleted in TG6002, enhancing selective replication in cancer cells [12]. In addition, insertion of the chimeric yeast *FCU1* gene enables the selective conversion of the prodrug 5-fluorocytosine (5-FC) into the cytotoxics 5-FU and 5-fluorouridine monophosphate (5-FUMP) [13], bypassing the natural resistance of tumour cells to 5-FU alone and reducing systemic toxicity [12]. Moreover, TG6002 induces an anti-tumour immune response involving CD8 T cells and tumour-infiltrating lymphocytes and myeloid cells [14].

TG6002 with 5-FC is a promising combination therapy for cancers that are sensitive to 5-FU. An open-label phase I dose-escalation trial of *i.v.* TG6002 plus 5-FC was initiated in 2018 (TG6002.02, NCT03724071) in patients with advanced gastrointestinal malignancies. Overall, the combination was well tolerated and no maximum tolerated dose (MTD) was observed. Preliminary results indicate effective biodistribution of TG6002 in tumour cells, associated with localised *FCU1* activity [15].

Critical to successful OV therapy is the delivery of virus to the tumour site. Various routes of administration have been investigated, predominantly *i.t*. and *i.v..* Intravenous administration is simple, relatively non-invasive and can achieve systemic delivery of virus to all vascularised tumours, although very high doses are required to achieve sufficient concentration at the tumour site, as the majority is redistributed throughout normal body organs. Intratumoural administration delivers the virus directly to the tumour; however, *i.t.* injection is limited to radiologically detectable, anatomically and technically injectable lesions, although abscopal effects have been reported at distant sites. For patients with liver-dominant cancers, an alternative route of delivery is via selective catheterisation of the hepatic artery; indeed, locoregional delivery of chemotherapy via the hepatic artery has been extensively studied [16,17] and was initially considered for administration of OVs in the early 2000s [18]. Intrahepatic artery (IHA) infusion of OVs has the potential to enhance delivery and distribution to multiple tumours across the liver, whilst limiting systemic chemotherapy toxicity. We describe the clinical and translational data from a dose-escalation study of TG6002 via hepatic artery administration plus oral 5-FC. The results show that IHA administration of an OV is clinically achievable, results in delivery of replication-competent virus to the tumour, expression and clinically-relevant activity of the *FCU1* transgene, peripheral activation of the immune system and potential ICD.

## METHODS

### Study design

TG6002.03 was an open-label, dose-escalation, 3 + 3 design, phase I study (Clinicaltrial.gov ID NCT04194034) conducted in three sites across the UK and France, in patients with unresectable CRC with liver metastases having progressed on or after standard chemotherapy including at least a fluoropyrimidine, oxaliplatin and irinotecan or, in the UK only, entering a period of clinical observation following discontinuation of chemotherapy. Patients received up to two cycles of TG6002 combined with oral 5-FC (Fig 1A). TG6002 was administered via the main hepatic artery, through a catheter inserted into the femoral artery under angiographic assessment, over 30 minutes at doses of 1×10^6^, 1×10^7^, 1×10^8^ and 1×10^9^ pfu. 5-FC was taken orally from day 5 to day 14 at a dose of 50 mg/kg four times daily. A second treatment cycle was to be administered from day 43 in the absence of disease progression or unacceptable toxicity. Dose escalation proceeded after review of safety data from each cohort by an independent safety review committee. Ethical approval was granted by Health Research Authority and Health and Care Research Wales, UK following approval by the following review boards: Medicines and Healthcare products Regulatory Agency UK, Clinical Trial Authorisation; 16280/0204/001-0001, Research Ethics Committee UK; 19/LO/1260; European Union Drug Regulating Authorities Clinical Trials; 2018-004103-39.

**Figure 1:**
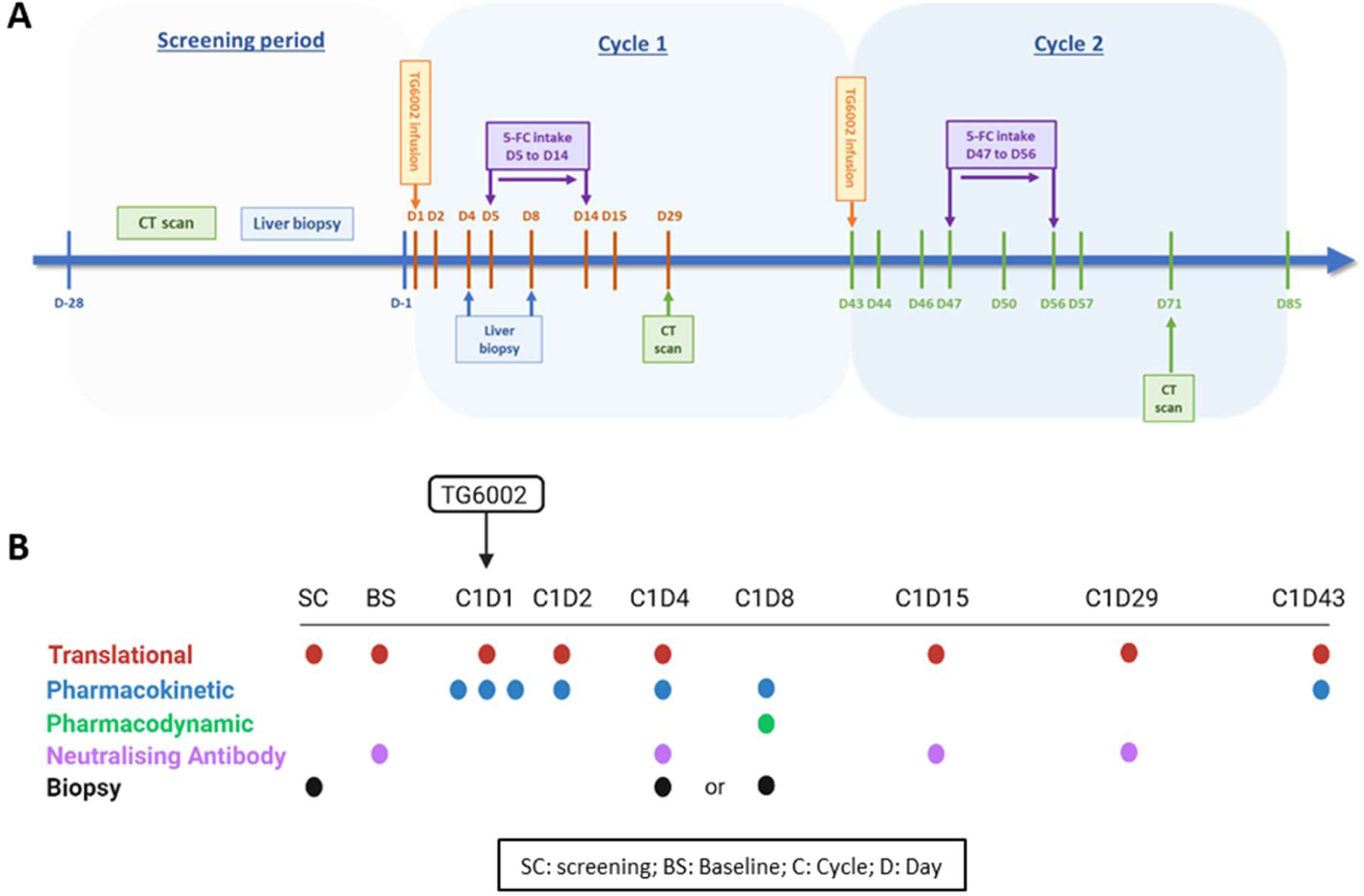
Trial schema and sampling schedule. (A) Trial schema depicting treatment schedule of two planned cycles of IHA TG6002 and oral 5-FC. (B) Patient blood and tissue samples were taken at various time points prior to and during treatment. Blood samples were specific for downstream analyses; translational (red), pharmacokinetic (blue), pharmacodynamic (green), NAb (purple) and tissue biopsies (black).

**Table 1:**
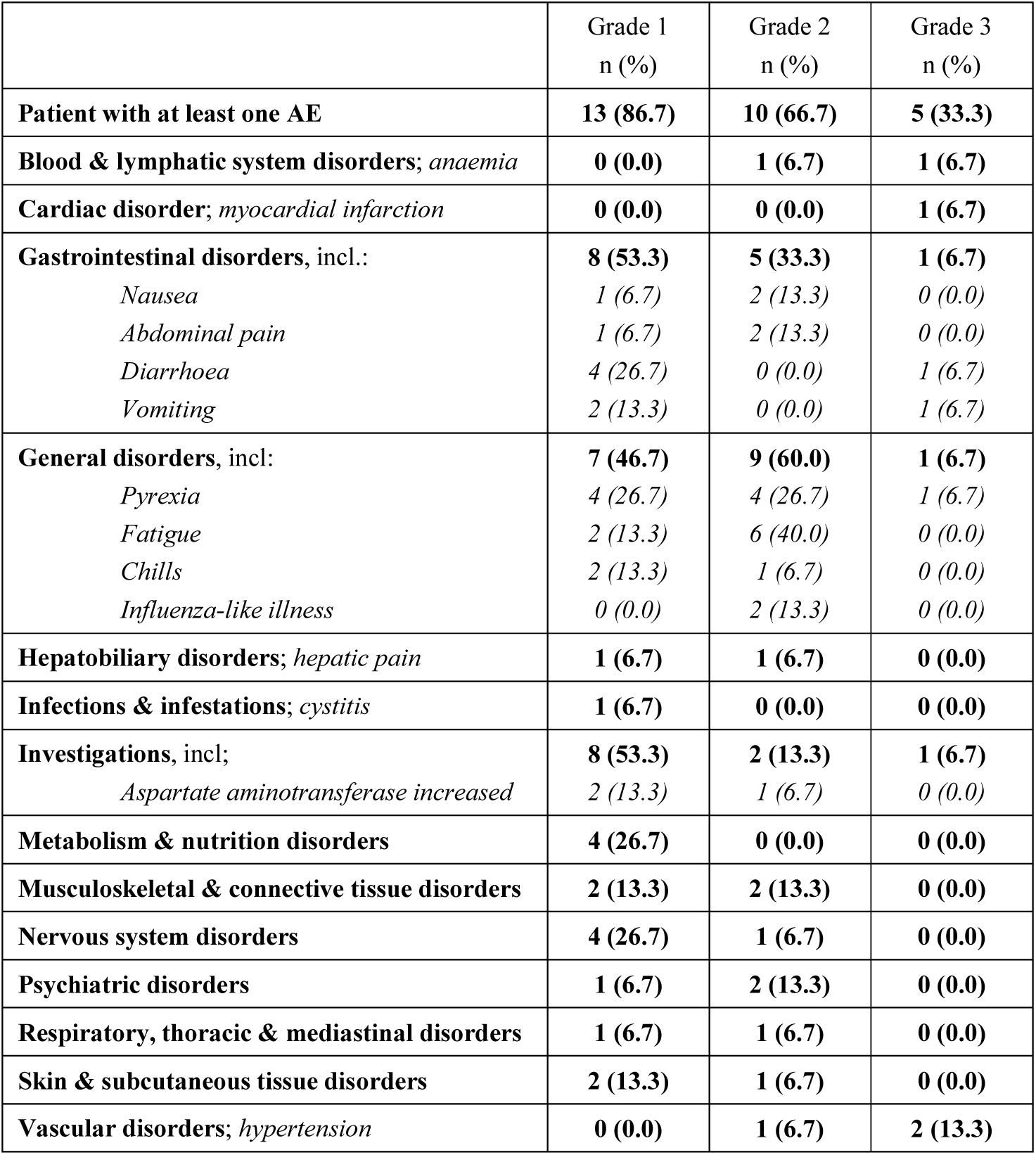
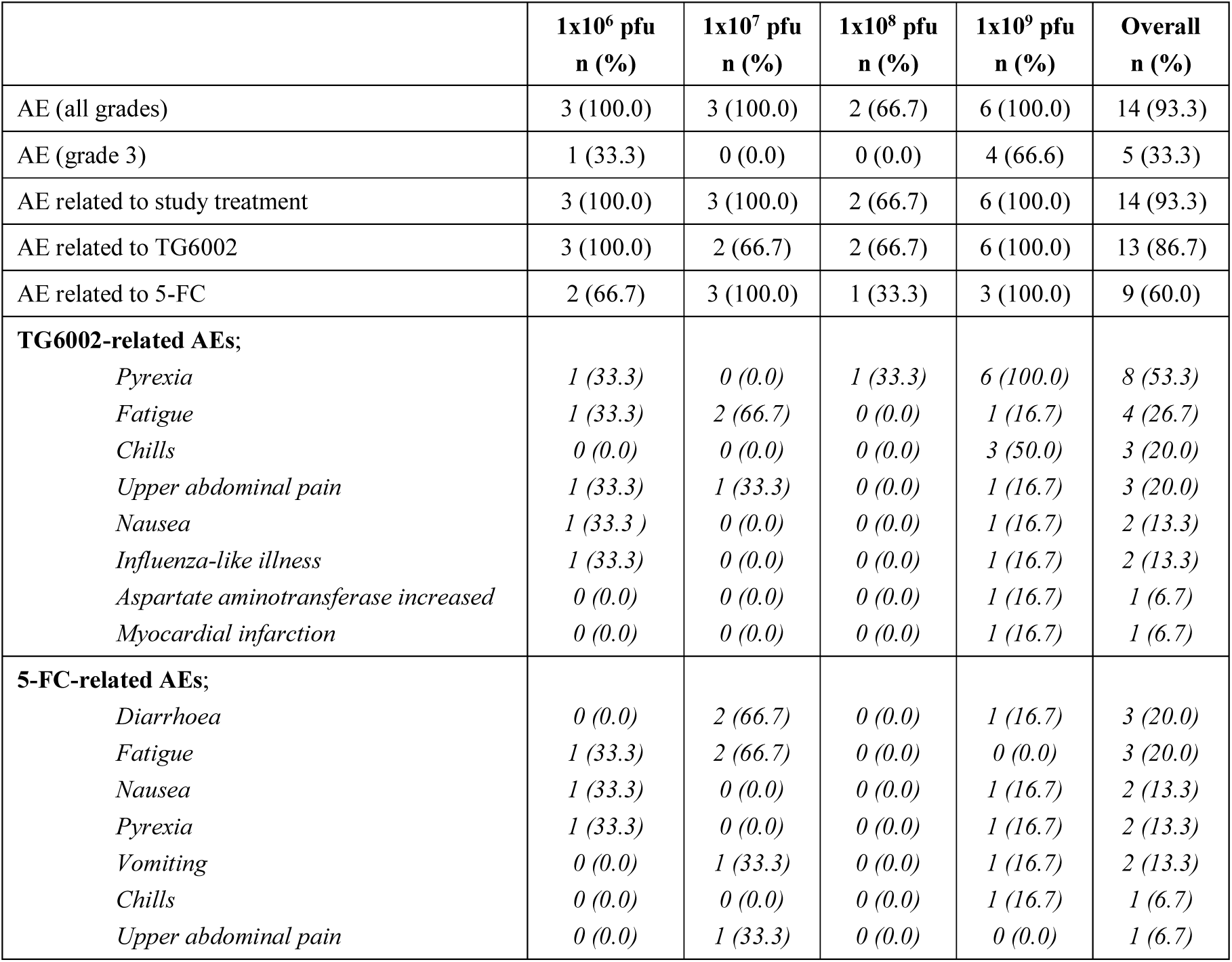
Adverse events summary. Adverse events summarised by relationship to TG6002 and 5-FC (A) and grade (B).

|  | Grade 1<br>n (%) | Grade 2<br>n (%) | Grade 3<br>n (%) |
| --- | --- | --- | --- |
| **Patient with at least one AE** | **13 (86.7)** | **10 (66.7)** | **5 (33.3)** |
| **Blood & lymphatic system disorders; anaemia** | **0 (0.0)** | **1 (6.7)** | **1 (6.7)** |
| **Cardiac disorder; myocardial infarction** | **0 (0.0)** | **0 (0.0)** | **1 (6.7)** |
| **Gastrointestinal disorders, incl.:** | **8 (53.3)** | **5 (33.3)** | **1 (6.7)** |
| *Nausea* | 1 (6.7) | 2 (13.3) | 0 (0.0) |
| *Abdominal pain* | 1 (6.7) | 2 (13.3) | 0 (0.0) |
| *Diarrhoea* | 4 (26.7) | 0 (0.0) | 1 (6.7) |
| *Vomiting* | 2 (13.3) | 0 (0.0) | 1 (6.7) |
| **General disorders, incl:** | **7 (46.7)** | **9 (60.0)** | **1 (6.7)** |
| *Pyrexia* | 4 (26.7) | 4 (26.7) | 1 (6.7) |
| *Fatigue* | 2 (13.3) | 6 (40.0) | 0 (0.0) |
| *Chills* | 2 (13.3) | 1 (6.7) | 0 (0.0) |
| *Influenza-like illness* | 0 (0.0) | 2 (13.3) | 0 (0.0) |
| **Hepatobiliary disorders; hepatic pain** | **1 (6.7)** | **1 (6.7)** | **0 (0.0)** |
| **Infections & infestations; cystitis** | **1 (6.7)** | **0 (0.0)** | **0 (0.0)** |
| **Investigations, incl;** | **8 (53.3)** | **2 (13.3)** | **1 (6.7)** |
| *Aspartate aminotransferase increased* | 2 (13.3) | 1 (6.7) | 0 (0.0) |
| **Metabolism & nutrition disorders** | **4 (26.7)** | **0 (0.0)** | **0 (0.0)** |
| **Musculoskeletal & connective tissue disorders** | **2 (13.3)** | **2 (13.3)** | **0 (0.0)** |
| **Nervous system disorders** | **4 (26.7)** | **1 (6.7)** | **0 (0.0)** |
| **Psychiatric disorders** | **1 (6.7)** | **2 (13.3)** | **0 (0.0)** |
| **Respiratory, thoracic & mediastinal disorders** | **1 (6.7)** | **1 (6.7)** | **0 (0.0)** |
| **Skin & subcutaneous tissue disorders** | **2 (13.3)** | **1 (6.7)** | **0 (0.0)** |
| **Vascular disorders; hypertension** | **0 (0.0)** | **1 (6.7)** | **2 (13.3)** |

|  | 1x10<sup>6</sup> pfu<br>n (%) | 1x10<sup>7</sup> pfu<br>n (%) | 1x10<sup>8</sup> pfu<br>n (%) | 1x10<sup>9</sup> pfu<br>n (%) | Overall<br>n (%) |
| --- | --- | --- | --- | --- | --- |
| AE (all grades) | 3 (100.0) | 3 (100.0) | 2 (66.7) | 6 (100.0) | 14 (93.3) |
| AE (grade 3) | 1 (33.3) | 0 (0.0) | 0 (0.0) | 4 (66.6) | 5 (33.3) |
| AE related to study treatment | 3 (100.0) | 3 (100.0) | 2 (66.7) | 6 (100.0) | 14 (93.3) |
| AE related to TG6002 | 3 (100.0) | 2 (66.7) | 2 (66.7) | 6 (100.0) | 13 (86.7) |
| AE related to 5-FC | 2 (66.7) | 3 (100.0) | 1 (33.3) | 3 (100.0) | 9 (60.0) |
| **TG6002-related AEs;** |  |  |  |  |  |
| *Pyrexia* | 1 (33.3) | 0 (0.0) | 1 (33.3) | 6 (100.0) | 8 (53.3) |
| *Fatigue* | 1 (33.3) | 2 (66.7) | 0 (0.0) | 1 (16.7) | 4 (26.7) |
| *Chills* | 0 (0.0) | 0 (0.0) | 0 (0.0) | 3 (50.0) | 3 (20.0) |
| *Upper abdominal pain* | 1 (33.3) | 1 (33.3) | 0 (0.0) | 1 (16.7) | 3 (20.0) |
| *Nausea* | 1 (33.3) | 0 (0.0) | 0 (0.0) | 1 (16.7) | 2 (13.3) |
| *Influenza-like illness* | 1 (33.3) | 0 (0.0) | 0 (0.0) | 1 (16.7) | 2 (13.3) |
| *Aspartate aminotransferase increased* | 0 (0.0) | 0 (0.0) | 0 (0.0) | 1 (16.7) | 1 (6.7) |
| *Myocardial infarction* | 0 (0.0) | 0 (0.0) | 0 (0.0) | 1 (16.7) | 1 (6.7) |
| **5-FC-related AEs;** |  |  |  |  |  |
| *Diarrhoea* | 0 (0.0) | 2 (66.7) | 0 (0.0) | 1 (16.7) | 3 (20.0) |
| *Fatigue* | 1 (33.3) | 2 (66.7) | 0 (0.0) | 0 (0.0) | 3 (20.0) |
| *Nausea* | 1 (33.3) | 0 (0.0) | 0 (0.0) | 1 (16.7) | 2 (13.3) |
| *Pyrexia* | 1 (33.3) | 0 (0.0) | 0 (0.0) | 1 (16.7) | 2 (13.3) |
| *Vomiting* | 0 (0.0) | 1 (33.3) | 0 (0.0) | 1 (16.7) | 2 (13.3) |
| *Chills* | 0 (0.0) | 0 (0.0) | 0 (0.0) | 1 (16.7) | 1 (6.7) |
| *Upper abdominal pain* | 0 (0.0) | 1 (33.3) | 0 (0.0) | 0 (0.0) | 1 (6.7) |

### Patient Samples

Blood and tissue samples were collected and processed using the Translational Cancer Immunotherapy Team quality-assured lab manual, which included standard operating procedures to regulate all processes. Peripheral blood was collected into K3EDTA or serum clot activator vacutainer tubes (both Scientific Laboratory Supplies) and processed within 2 hrs of venepuncture, or as soon as possible thereafter. Tumour biopsies collected on day 4 or day 8 were placed in formalin or RNALater (both ThermoFisher) for IHC or PCR analyses, respectively. All sample collection time points are shown in Fig 1B.

### Isolation of peripheral blood mononuclear cells (PBMCs), plasma and serum from whole blood

Serum clot activator tubes were left for a minimum of 30 mins post-venepuncture. All blood collection tubes were then centrifuged for 10 mins at 2000 g. Plasma and serum aliquots from the upper layers were stored at -80 °C. PBMCs were isolated by density-gradient centrifugation over lymphoprep™ (StemCell Technologies) as per manufacturer’s instructions. Cells were frozen at 1×10^7^/ml in 40 % (*v/v*) Roswell Park Memorial Institute (RPMI) medium containing 5 mM L-Glutamine and 1 mM sodium pyruvate (all Sigma), plus 50 % (*v/v*) pooled human serum (SeraLab) and 10 % (*v/v*) dimethyl sulphoxide (DMSO; Sigma). PBMCs were stored in liquid nitrogen.

### Detection of virus in tumour biopsies and plasma

qPCR: DNA was extracted from tumour biopsies using DNeasy Blood and Tissue Kits (Qiagen).

Primers (forward 5’-CGATGATGGAGTAATAAGTGGTAGGA-3’ and reverse 5’-CACCGACCGATGATAAGATTTG-3’) (Integrated DNA Technologies) were used to detect for the presence of TG6002.

<u>RT-qPCR</u>: was performed on tumour biopsies and plasma for detection of viral early *D7R*, viral late *A10L* and *FCU1* transcripts. RNA was extracted using RNeasy plus Mini Kits (Qiagen). Remaining viral and cellular DNA in samples were digested with Turbo DNAse (ThermoFisher). Primers and probes for D7R (forward TTTAGCGATTCAAAGTACTGCTTTTT, reverse GCAGTGACTTCGCTGCCATT, probe FAM-CGAAATGGTAATGCGTATGA); A10L (forward CTTCATACTCGCGATCCTCAAA, reverse TCGCCAACAGGTTAAAGAAATTAA, probe ABY-TGGCGCTTCCAAACGTGCAATTT); FCU1 (forward TCGTGGTCACAACATGAGATTTC, reverse TCTAATCTCCCACAGTTTTCCAAAG, probe ABY-TCCGCCACACTACATGGTGAGATCTCC) were used. Detection of TG6002 viral genome in plasma was performed by Charles River Laboratories Evreux using in-house methods. All PCR data was acquired on Applied Biosystems QuantStudio™ 5 Real Time PCR Systems (ThermoFisher) and analysed using QuantStudio 3D AnalysisSuite Cloud (ThermoFisher). The presence of the virus in tumour biopsies was considered positive by RT-qPCR if at least one of the viral mRNAs (*D7R*, *A10L* or *FCU1*) was detected. RNase-free water was used as negative controls.

<u>Plaque assays:</u> were performed by Transgene, France, using in-house methods. Briefly, tissue biopsies were sonicated for 15 seconds at room temperature before incubation with a permissive cell line, Vero. Confluent monolayers were incubated with tumour samples for 30 mins prior to incubation at 37 °C under 1 % agarose for 3 days. Positive infection was determined by the presence of viral plaques.

<u>IHC:</u> for viral protein was performed by Cerba Research, France. A polyclonal anti-vaccinia virus antibody (Tebu-bio) was used to detect virions; negative control was secondary antibody alone. DAB-HRP peroxidase was used to visualise virus-positive cells. Data from all methods are expressed as positive or negative/below level of detection. Biopsies from two patients (13 and 15) were not available.

### 5-FU concentrations in serum and tumour tissue

Quantification of 5-FC, 5-FU and 5-fluoro-β-alanine (F-BAL) levels was performed using liquid chromatography coupled with high-resolution mass spectrometry (Hospices Civils de Lyon, France) as described previously [19].

Serum was evaluated on day 8-post TG6002. Additionally, 5-FU concentrations were measured in available tumour biopsies on day 8. F-BAL was not measured in cohort 1 serum samples or tumour biopsies due to sample insufficiency.

### Calreticulin ELISA

Patient plasma was analysed for calreticulin (CRT) by ELISA (ThermoFisher) as per manufacturer’s instructions. Data is expressed as mean plasma concentration (ng/ml) ± SEM, calculated using a standard curve. Statistical significance was determined using paired T tests between sample time points (* P<0.05); n=14 patients, dependent on sample availability.

### mRNA expression analysis of patient PBMCs

mRNA sequencing was performed by Novogene as per validated methods. After PCR, gene expression level was calculated by the number of mapped reads. Statistically significant differentially-expressed genes (ssDEGs) were defined as ± >2 log2FoldChange of post-treatment samples compared to baseline with an associated Padj <0.05. Data from six patients, across three cohorts, are shown for ssDEGs for C1D2 and C1D15. Padj values were transformed into –log10(Padj) values, which were plotted against log2FoldChange values in volcano plots. Volcano plots depict all DEGs, not just ssDEGs, which are upregulated, downregulated or unchanged for three patients at C1D2 and C1D15 compared to BS. ssDEG lists were analysed using the Search Tool for Retrieval of Interacting Genes/Proteins (STRING) database (http://string-db.org) [20] to identify potential interactions between the genes and their reported biological function(s). Interaction Confidence Scores (ICS) were assigned to each protein association and ranked from 0 to 1, where 1 is most likely to be accurate and 0 least likely to be correct. An ICS of 0.5 indicates that every second interaction may be a false positive, therefore, only ssDEGs with an ICS score of >0.5 were used for visualisation and analysis. Gene Ontology (GO) biological functions were assessed at the

ssDEG level, where common genes were detected across multiple patients; highlighted green in the volcano plots. These nine commonly expressed ssDEGs were analysed independently in the STRING database to identify highly responsive signaling pathways following treatment.

### Immunophenotyping

PBMCs were analysed for specific activation/immune checkpoint molecules. Briefly, PBMCs were stained for CD3 (HIT3a, FITC), CD4 (RPA-T4, APC-H7), CD8 (RPA-T8, Alexa 700), CD56 (B159, PE-Cy7), CD19 (SJ25C1, APC-H7), CD14 (M5E2, FITC), CD69 (FN50, APC), PD-L1 (MIH1, PE-CF594), PD-1 (MIH4, PE), TIM-3 (7D3, BV786), OX40 (L106, BV421), CD40 (5C3, BUV395), CD25 (M-A251, BB700) (all BD), plus

CD40L (24-31, BV786) and CTLA-4 (BNI3, BV605) (both Biolegend). Fluorescence minus one (FMO) were used as negative controls. Data was acquired on a CytoFlex LX and analysed using CytExpert software (both Beckman Coulter). Positive expression of markers was used to calculate fold-change differences ± SEM in expression from baseline samples. Paired T tests were used to determine statistical significance between samples (* P<0.05), n=9 patients, dependent on sample availability.

### IHC for Programmed death-ligand 1 (PD-L1)

Formalin-fixed paraffin-embedded (FFPE) tissue biopsies were stained with rabbit anti-human PD-L1 antibody (Abcam; 1:500) and ImmPRESS-HRP anti-rabbit IgG (peroxidase) secondary antibody (Vector). Positive staining was visualised using ImmPACT DAB Peroxidase (HRP) Substrate kit (Vector). Control sections contained no primary antibody. Digital images were acquired at x20 magnification and quantified using QuPath software [21]. Data is expressed as cells positive for PD-L1 per mm^2^ for screening (black bars) and day 8 biopsies (hatched bars) for n=7 patients.

### ELISpot

Briefly, triplicates of 1×10^5^/well PBMCs were incubated in the presence of either 2 µg/ml of overlapping peptide pools for carcinoembryonic antigen (CEA) or cytomegalovirus/Epstein-Barr virus/influenza (CEF; positive control) (both Cambridge Biosciences). Negative control was media alone; 10 pfu/cell TG6002 was used to assess response to treatment. Interferon (IFN)-γ secretion from activated T cells was detected using a matched paired antibody kit (MabTech). Spot-forming units (SFU) were visualised using BCIP/NBT substrate (MabTech). Images were captured and quantified using an S6 FluoroSpot analyser (Cellular Technology Limited). Data are presented as mean fold-change SFU per well ± SEM for post-treatment samples compared to baseline, for n=6 patients, dependent on sample availability.

### T cell receptor (TCR)-β sequencing

TCRβ sequencing was performed by Adaptive Technologies using a ‘survey’ resolution to generate data from productive rearrangements only, which were exported from the immunoSEQ Analyser for further analysis. Complementary-determining region 3 (CDR3) sequences were input into the McPAS TCR database [22] and matched to known human TCR sequences. TCRs in each patient sample that matched to known cancer or neoantigen epitopes were identified; these were counted and their total productive frequency calculated. Data is expressed as number of TCRs matching cancer antigens/neoantigens (*x*-axis) against productive frequency of TCRs matching cancer antigens/neoantigens (*y*-axis) for all available PBMCs samples.

## RESULTS

### Patient characteristics

In total, 20 patients were screened; of these, 15 patients were entered into the study across three sites (Supplementary Table 1) and received at least one infusion of TG6002. Mean age was 61 years (range 37 to 78 years), comprising eleven males and four females. Patients had mismatch repair proficient cancers. Included patients had either progressed or were intolerant to both Oxaliplatin and Irinotecan-based chemotherapy or were undergoing a period of observation following a course of chemotherapy. Primary tumour was in the colon in eleven patients and the rectum in four patients. Mean time from initial diagnosis to trial entry was 36.5 months (range 8.1 to 89.8 months) and the patients had received a mean of 3.3 prior lines of antineoplastic therapy (range 1 to 7), including adjuvant lines. Thirteen patients completed the trial; two patients prematurely withdrew due to disease progression or death.

### Patient exposure

Of the 15 patients entered, three were treated in each of cohorts 1, 2 and 3, with six treated in cohort 4 (Supplementary Table 2); associated trial IDs and manuscript IDs are depicted for clarity: these patient IDs are not known to anyone outside of the research team.

Fourteen patients received a single cycle of treatment and one received two cycles; each cycle being a single dose of TG6002 via IHA infusion on day 1 followed by 10 days of oral 5-FC on days 5-14 (Fig 1A). All infusions were fully administered. In 13 patients, the whole liver was perfused while two patients had partial liver perfusion due to anatomical considerations. One patient did not receive 5-FC having withdrawn from the trial on day 1 after the TG6002 infusion and before receiving 5-FC. Thirteen of the remaining 14 patients received their complete 10-day course of 5-FC but one patient discontinued 5-FC after nine days.

### Safety data

Overall, 14 patients (93.3 %) experienced at least one study treatment-related adverse event (AE), of whom thirteen (86.7 %) experienced at least one AE related to TG6002 and nine (60.0 %) experienced at least one AE related to 5-FC (Tables 1A and B).

Eight grade 3 AEs were observed in five patients, including myocardial infarction (MI), diarrhea, vomiting, pyrexia and increased aspartate aminotransferase related to study treatment (Tables 1A and B), plus hypertension and anaemia unrelated to treatment. Overall, no AEs greater than grade 3 were reported. Grade 3 TG6002-related AEs included pyrexia and MI in one cohort 4 patient, which constituted a dose-limiting toxicity (DLT) (Tables 1A and B). This patient

acquired an asymptomatic COVID-19 infection near the time of TG6002 infusion, precipitating a supraventricular tachycardia. Four hours following TG6002 infusion, the patient had a fever and tachycardia that peaked at 40.1 °C and 148 bpm. On day 2, an electrocardiogram (ECG) showed negative T-waves and troponin I level was increased at 1555 pg/l (N<45). A coronary angiography was performed on day 3 showing a monotruncular lesion of the anterior interventricular artery leading to coronary stent insertion and successful revascularisation. No cases of vesicular or pustular skin or mucosal lesions were reported following TG6002 infusion. 5-FC-related grade 3 AEs included diarrhea and vomiting in one cohort 4 patient. Aspartate aminotransferase increase occurred in three patients; grade 1 in two patients, with one case related to TG6002, and grade 2 in one patient. Hypertension was reported for three patients; grade 2 in a cohort 2 patient and grade 3 in a patient each in cohorts 1 and 4. Despite being assessed as not related to study treatment, this resulted from antihypertensives being withdrawn during TG6002 administration.

### Trial endpoints and objective efficacy data

No patients had an objective response based on a 10-week disease control rate according to RECIST v1.1. The primary objective of maximum feasible dose (MFD) was 1×10^9^; MTD was not reached. Secondary objectives of safety and tolerability were achieved; in addition, viral shedding was not evident in saliva, urine or faeces. Median progression-free survival was 1.05 months, with a range of 0.0 to 2.3 (where 0.0 relates to the MI reported previously), which is very short due to the timing of the CT scan 4 weeks post-TG6002 infusion ahead of the planned second TG6002 infusion. Median overall survival was 5.4 months (range 1.6 to 22.3).

### Detection of TG6002 and FCU1 trangene activity in tumour biopsy

Blood and tissue samples for translational analyses were taken as outlined in the trial schedule (Fig 1B). As only one patient received a second cycle of treatment, the translational assays for all patients were performed on samples obtained during the first cycle only. Tumour biopsies obtained at screening and post-treatment (day 4 or day 8) were examined for the presence of TG6002 virus or for activity of the viral *FCU1* transgene (Fig 2). Viral DNA by qPCR was detected in five out of 13 evaluable tumour samples, predominantly in biopsies from patients who received a higher viral dose than patients in earlier cohorts, both at days 4 and 8 (Fig 2A). Furthermore, virus was detected by RT-qPCR in four of nine patients with suitable biospecimens. Plaque assays indicated live replicating TG6002 in two of nine tumours. Viral protein was detected by IHC in three of six evaluable biopsies; one at day 4 and two at day 8, with representative examples shown in Fig 2B. The active conversion of 5-FC to its metabolite, 5-FU, was detected in three of six evaluable post-treatment biopsies, predominantly in tumours of patients in later cohorts, suggesting a higher virus dose is required for TG6002 activity within tumour. One patient in cohort 2 (one out of two available biopsies) had 16.2 pg of 5-FU/mg tumour tissue, whereas two patients in cohort 4 (two out of three available biopsies) had 35 and 29 pg of 5-FU/mg tumour tissue. Overall, there was evidence of virus infection and viral replication in ten of 13 patients’ tumours.

**Figure 2:**
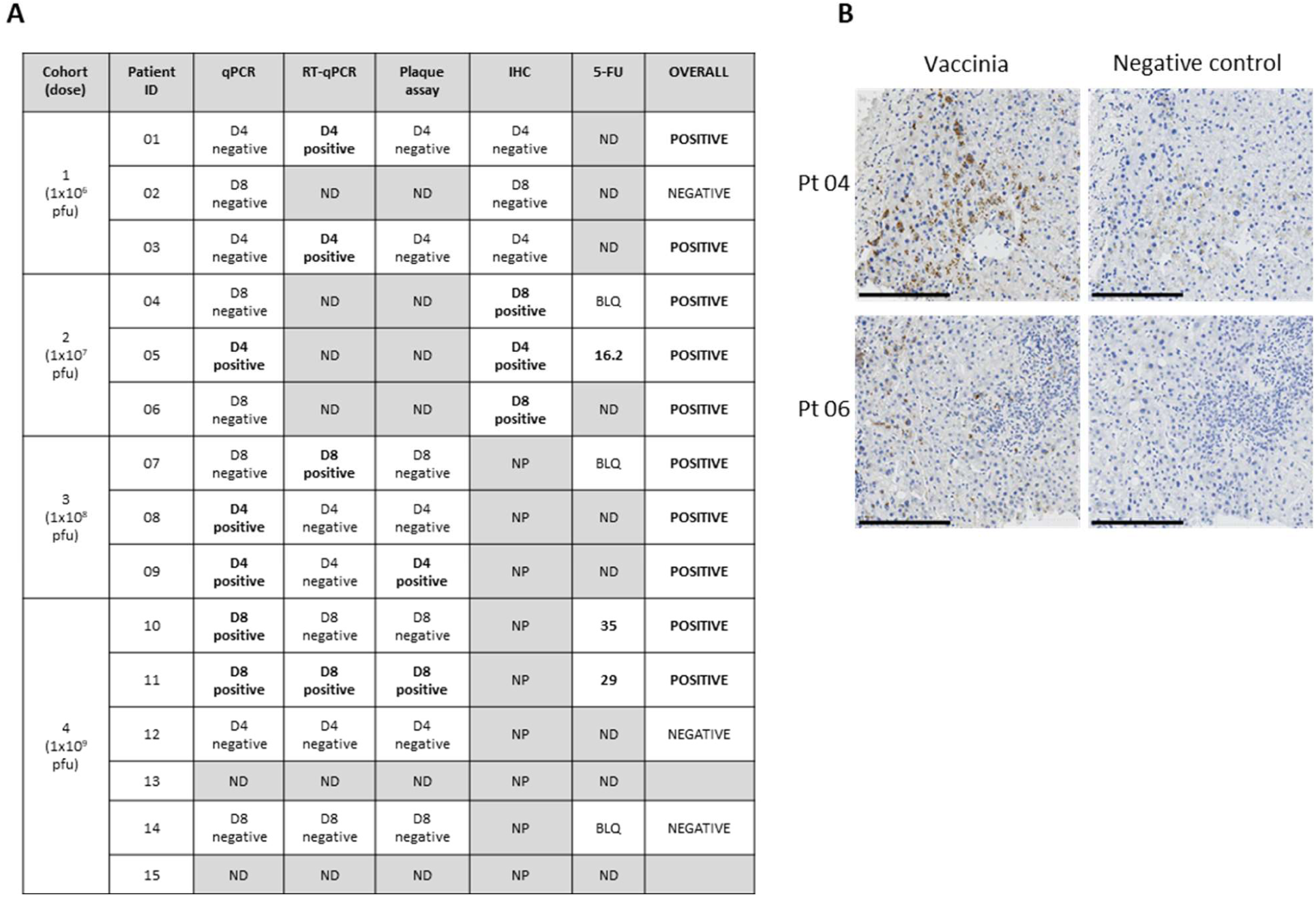
Detection of TG6002 in patient tumor biopsies. (A) Table summary of data collated from assays to detect the presence of TG6002 in tumor biopsies: qPCR and RT-qPCR to detect viral nucleic acids, plaque assay to detect replication-competent virus, IHC to determine the presence of virus protein and 5-FU (pg/mg) to demonstrate transgene activity. Samples are designated as either positive or negative/below the limit of detection for each assay. ND: not done due to insufficient sample; NP: not planned; BLQ: below limit of quantification; D4: day 4; D8: day 8. (B) Representative examples of positively-stained tumour cells by IHC are shown for patients 04 (day 4) and 06 (day 8) (left panel; brown DAB), negative control (right panel; secondary antibody alone). Scale bars represent 200 µm.

### Detection of TG6002 and FCU1 trangene activity in plasma

Despite detection in tumours, TG6002 was not found in the vast majority of plasma samples from cohorts 1-3, with the exception of one patient in cohort 2 and one in cohort 3, both at day 8, indicating active virus replication (Fig 3A). In contrast, in the highest dose cohort, TG6002 was detected in plasma 30 minutes post-infusion in five of six patients, followed by undetectable levels indicating rapid clearance. A rebound of circulating TG6002 was observed in one patient at day 4 and three patients at day 8, again indicating active virus replication. Titres of NAb against TG6002 significantly increased following treatment in all patients (P<0.05), with a trend for higher titres in the highest dose cohort (Fig 3B).

**Figure 3:**
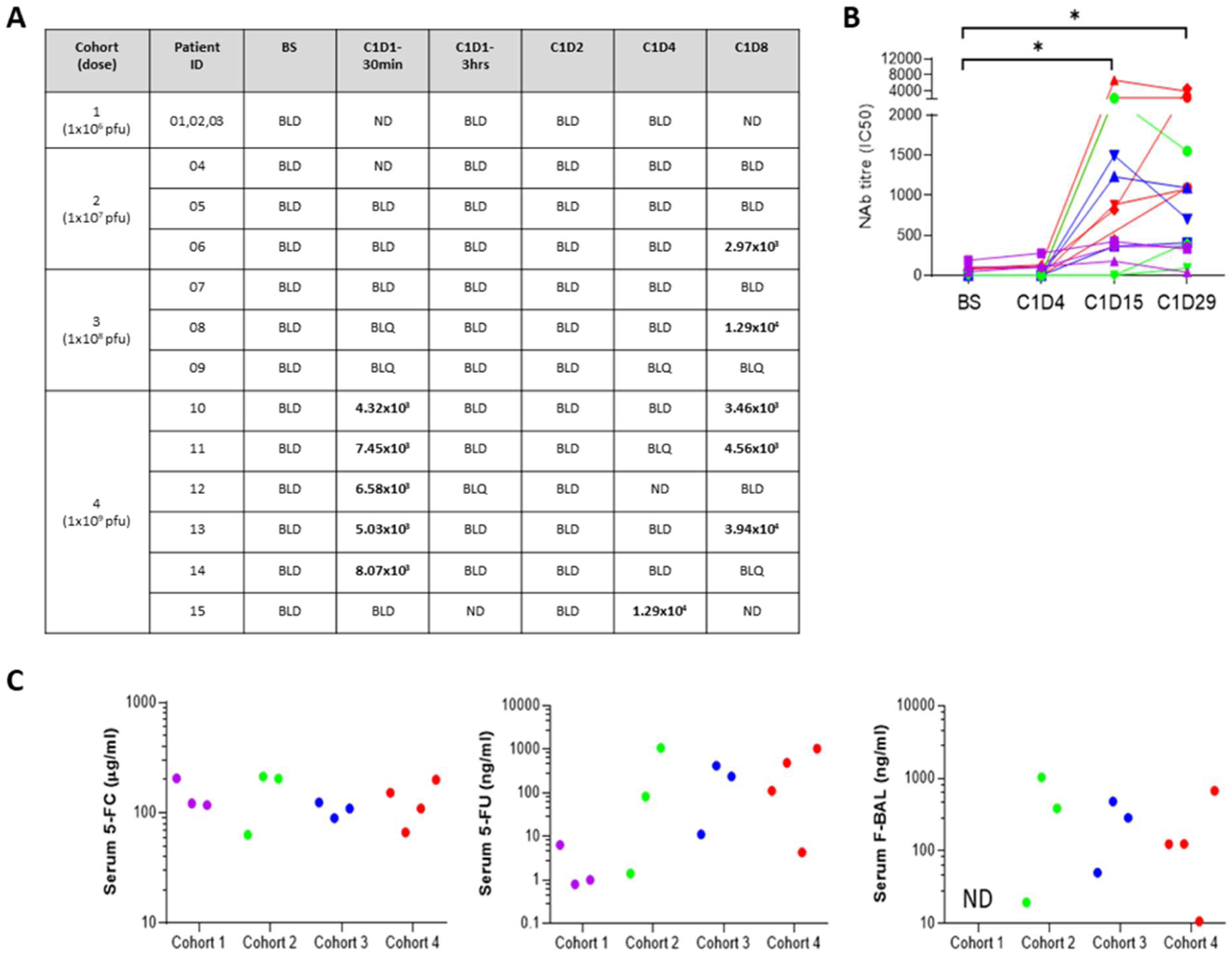
Detection of TG6002, NAb, 5-FC, 5-FU and F-BAL in plasma following treatment. (A) Plasma samples were analysed by qPCR to detect TG6002 following treatment. Values stated are number of copies/ml (c/ml); BLD: below limit of detection; BLQ: below limit of quantification; ND: not done due to insufficient sample. (B) Serum NAb titres (IC50) for N=15 patients, sample availability-dependant. * P<0.05, paired T test. (C) Serum 5-FC, 5-FU and F-BAL concentrations at D8 following treatment (N=13 patients), sample availability-dependent. Cohort 1: purple; cohort 2: green; cohort 3: blue; cohort 4: red.

Serum levels of 5-FC, 5-FU and the catabolite F-BAL was measured eight days after exposure to TG6002. Whereas serum 5-FC concentrations were comparable across all cohorts, patients who received higher virus doses had higher levels of circulating 5-FU than those in cohort 1, indicating replication of TG6002 (Fig 3C). F-BAL was detected in plasma from all patients from cohorts 2-4, where samples were available. Similar to the tumour data, there was evidence of viral presence/replication in serum in all evaluable patients.

### Host response to TG6002/5-FC

The peripheral immune response to IHA infusion of TG6002 was assessed using serial blood samples from patients. Calreticulin was investigated as an indicator of ICD following treatment. CRT concentration in patient plasma significantly increased following TG6002 infusion (Fig 4A); a peak was detected at 6 hours post-treatment (P<0.05), which remained higher than pre-treatment levels at day 2 (P<0.05), potentially indicating a peak in ICD.

**Figure 4:**
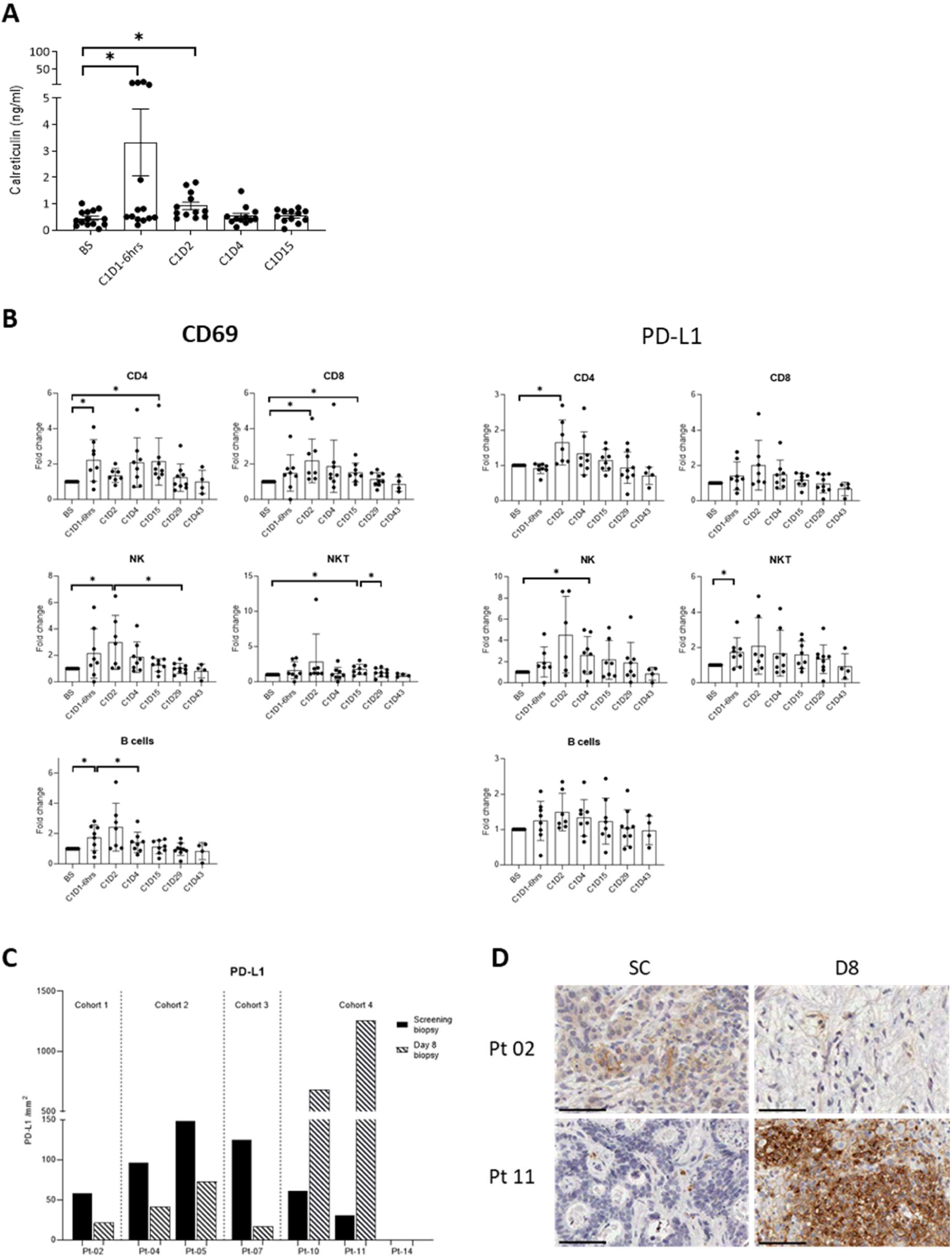
Activation of patient peripheral immune responses following treatment. (A) Calreticulin concentration (ng/ml) in patient plasma measured by ELISA. Data is expressed as the mean ± SEM; N=14 patients, sample availability-dependent. * P<0.05, paired T test. (B) Immunophenotyping of patient PBMCs for expression of CD69 & PD-L1. Relevant cell populations are depicted for each plot. Data is expressed as the mean fold-change ± SEM; N=9 patients. * P<0.05, paired T test. (C) Day 8 tumour PD-L1 expression by IHC is expressed as positive cells per mm^2^ in screening (black bars) and day 8 post-infusion (hatched bars) biopsies, with representative examples of patients from cohort 1 and 4 (D); screening, left panel; day 8, right panel. Positive staining by DAB (brown); scale bars represent 50 µm.

Immunophenotyping of PBMCs revealed CD69 upregulation, an early activation marker, 6-24 hours post-infusion on cell populations including CD4+ and CD8+ T cells, NK cells, NKT cells and B cells (Fig 4B). A rise in PD-L1 expression was also observed, as exemplified on NK cells (Fig 4B) alongside other immune checkpoint molecules, such as programmed cell death protein 1 (PD-1), T-cell immunoglobulin and mucin-domain containing-3 (TIM-3) and OX40 (Supplementary Fig 1A). In addition, elevation in CD40L on T cells and NK(T) cells (Supplementary Fig 1B) and an associated increase in its receptor (CD40) on both monocytes and B cells (Supplementary Fig 1C) was found, indicating enhanced capacity for the maturation of antigen presenting cells. In contrast, a decrease in both CD25 and cytotoxic T-lymphocyte associated protein 4 (CTLA-4) on the surface of T cells was apparent, which appeared to be prolonged over time, indicating reduced regulatory T cell functions (Supplementary Fig 1D).

Immunohistochemistry on tumour biopsies (Fig 4C) sampled pre- and post-infusion showed low-level PD-L1 expression at baseline and an apparent reduction in the level of PD-L1 in the tumour by day 8 in patients receiving a lower virus dose (cohorts 1-3). However, with the highest dose (cohort 4), there was a substantial increase in expression of PD-L1 following virus infusion, reflecting the PBMC data. Representative examples for patients in cohorts 1 and 4 are shown, indicating the extent of cells positive for PD-L1 in patients who received the higher viral load (Fig 4D).

mRNA sequencing was used to characterise the effects of TG6002/5-FC treatment at the transcriptional level (Fig 5). A greater number of ssDEGs were increased at day 2 in PBMCs from patients in cohorts 2 and 3 compared to patients in cohort 1 (Fig 5A). Patients from cohort 1, who did not show elevated ssDEGs by day 2, had a greater number of ssDEGs at day 15, potentially indicating a delayed response to replicating virus or to 5-FC and its metabolites, including 5-FU. Volcano plots (Fig 5B) show the pattern of all DEGs in three patients. Nine commonly expressed genes were significantly upregulated in response to TG6002 in three patients, all of which are involved in immune-related signaling pathways. Whereas these specific ssDEGs were evident by day 2 in Pt-07 (cohort 3), an increase in expression was only apparent in patients from cohort 1 (Pt-01 and Pt-03) by day 15. GO analysis of all upregulated ssDEGs generated cluster plots depicting pathways in which the DEGs are highly involved (Figs 5C, D and E). Cluster analysis of the nine commonly expressed ssDEGs in the three specified patients (Fig 5C) revealed a number of pathways highly relevant to immune responses, type I interferon signaling and more specifically, response to virus (Fig 5D). Each pathway identified had a significant proportion of the nine ssDEGs involved, as indicated. More widely, predominant clustering of all ssDEGs highlight several immune-related signaling pathways.

**Figure 5:**
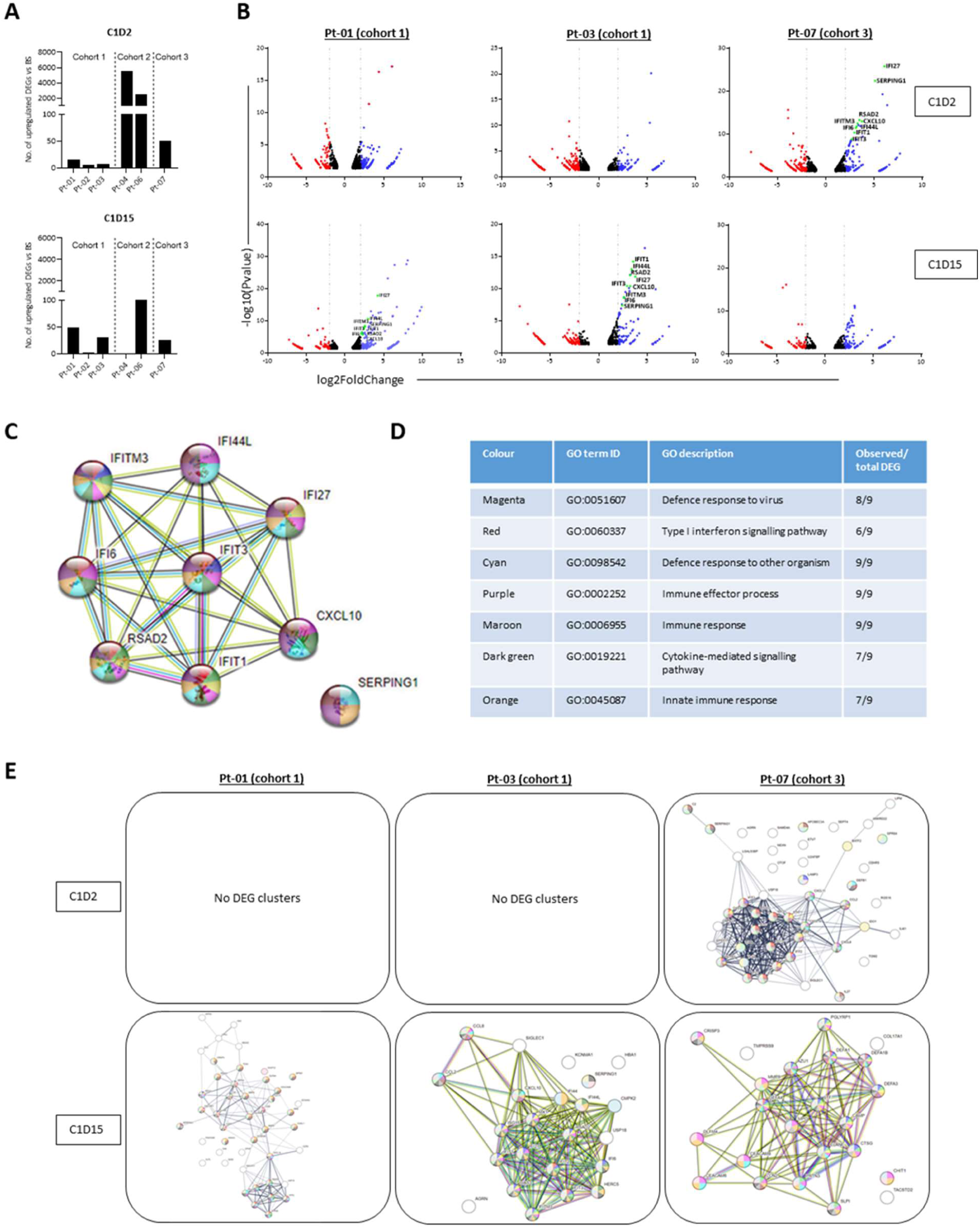
mRNA sequencing of patient PBMCs reveals clustering of DEGs involved in immune activation & response pathways to treatment. (A) The number of upregulated ssDEGs in the post-treatment samples relative to pre-treatment controls are shown for days 2 & 15 post-infusion. (B) Volcano plots for three patients which highlight nine commonly expressed ssDEGs (red: downregulated DEGs; blue: upregulated DEGs; black: non-significant changes to DEGs; green: specific highlighted ssDEGs). (C) Cluster plot of the nine commonly expressed ssDEGs in three patients which correspond to (D) immune-related GO pathways. (E) GO analysis showing cluster plots of all upregulated ssDEGs in three patients at days 2 & 15. DEGs are defined as up/downregulated from baseline, Padj<0.05; ssDEGs are defined as ± >2 log2FoldChange, Padj<0.05.

Pathway clustering was apparent by day 2 in the cohort 3 patient in comparison to day 15 in cohort 1 patients (Fig 5E). A greater extent of clustering mirrors both the enhanced number of ssDEGs and their earlier appearance following TG6002 infusion, as previously observed (Fig 5A).

The adaptive T cell response to virus infusion was examined by ELISpot assay against a tumour-associated antigen (TAA; CEA) and TG6002 (Figs 6A and B). CEA-specific T cell responses were detected by day 4, likely indicating enhanced activation of pre-existing CEA-specific T cell clones. In contrast, the appearance of TG6002-specific T cells occurred later, at day 15 (Fig 6A). Furthermore, only patients in the later cohorts (2 and 3) elicited TG6002-specific T cell responses, presumably due to a higher virus load. Representative wells showing IFNγ responses to CEA and TG6002 are depicted for two patients (Fig 6B).

**Figure 6:**
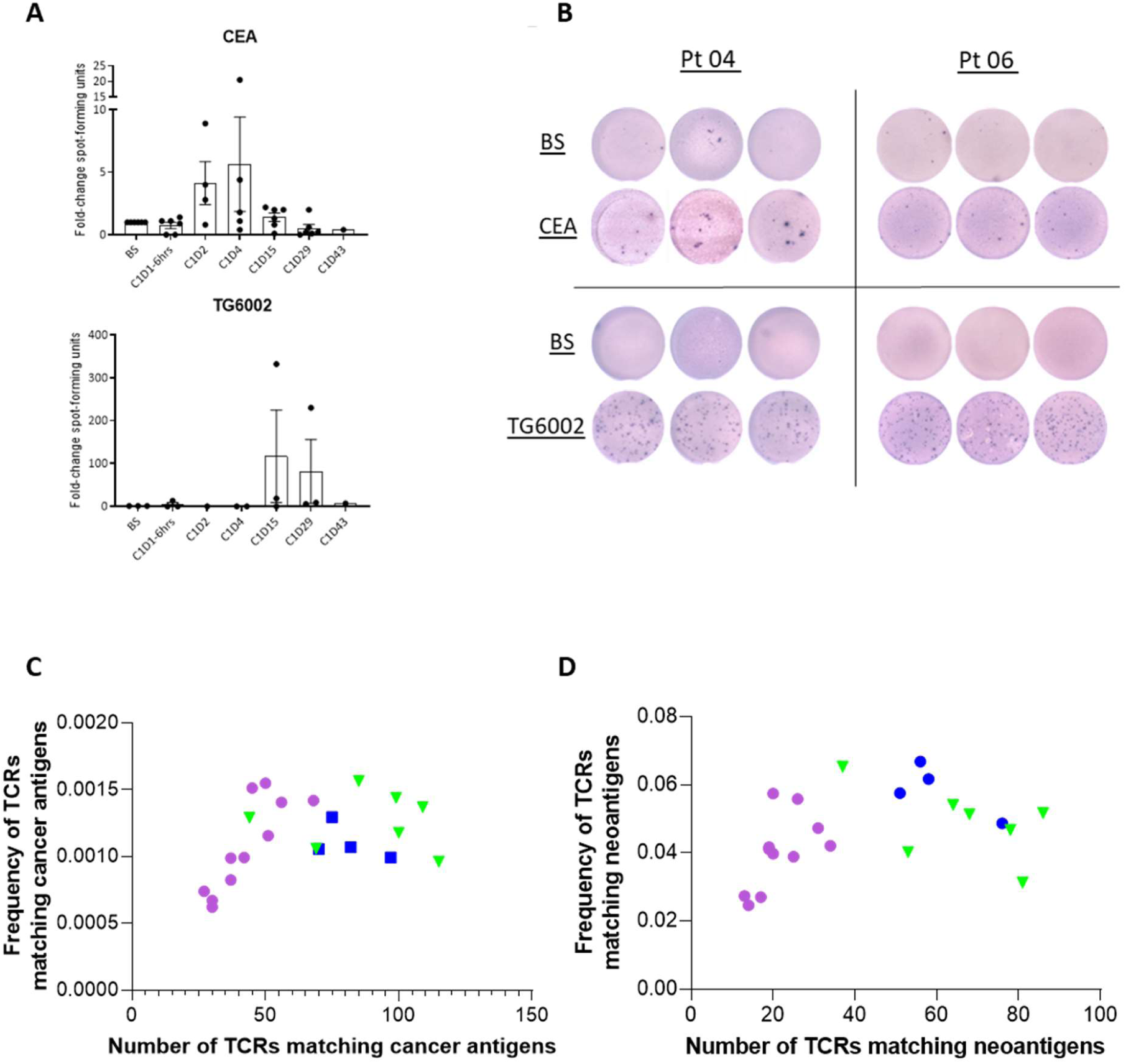
Adaptive T cell responses to treatment. T cell function was assessed by ELISpot assay, which showed an IFNγ-release response to CEA & TG6002. (A) Data is expressed as the mean fold-change ± SEM SFU of post-treatment samples compared to baseline. N=6 patients, sample availability-dependant. (B) Representative examples of SFU from two patients showing BS & post-treatment samples, in triplicate, depicting responses to CEA & TG6002. (C&D) TCRβ sequencing examined the T cell clonal response to TG6002 infusion. CDR3 sequences were matched to known TCRs using the McPAS database. TCRs matched to cancer (C) and neoantigen epitopes (D) are shown; data is expressed as number of total matched TCRs (*x*-axis) vs sum of productive frequencies of matched TCRs (*y*-axis). Cohort 1; purple; cohort 2: green; cohort 3: blue.

TCRβ sequencing of patient tumour and PBMCs at all available timepoints was also performed. T cell clonal response to treatment revealed CDR3 sequences matched to cancer antigens (Fig 6C) and specifically to neoantigens (Fig 6D), which showed greater frequencies in later cohorts treated with higher viral doses than in earlier lower-dose cohorts.

## DISCUSSION

In total, 15 patients received an IHA infusion of TG6002 plus oral 5-FC as part of a dose-escalation phase I study highlighting the feasibility of this locoregional route of oncolytic virus delivery in the treatment of liver tumours. Intrahepatic artery delivery of TG6002 was clinically feasible and safe, with the MTD not reached. Dose escalation proceeded as per the protocol, with six patients receiving the highest intended dose of 1×10^9^ pfu. Only one patient, in cohort 4, had a DLT that consisted of the aforementioned myocardial infarction. Disappointingly, no patients experienced clinical or radiological tumour responses, with almost all patients showing continued disease progression one month-post TG6002 delivery. The reasons for this could include the heavily pre-treated population; patients in this trial had all exhausted standard chemotherapy options, meaning that their cancers are 5-FU-resistant. Patients in this trial were not selected on the basis of mismatch repair (MMR) tumour status and no tumours were known to be MMR-deficient within the recruited cohort. It is likely that all or the majority of recruited patient tumours were MMR-proficient and relatively resistant to immunotherapy so less likely to benefit from OV therapy.

Successful delivery of TG6002 to tumour lesions was achieved via the IHA route. Viral persistence in tumour biospies sampled post-treatment was evident from a number of analyses, including qPCR, RT-qPCR, plaque assay, IHC and transgene activity, with the vast majority of patients exhibiting positive detection by one or more methods, despite only limited tissue from a core biopsy being available for analysis. Neutralising antibodies against TG6002 developed at low levels following infusion, reaching peak titres by day 15/29, indicating a humoral immune response to IHA delivery. It is unknown how the presence of low-level NAb might impact the repeated delivery of TG6002 via IHA; further investigation of locoregional oncolytic vaccinia virus therapy for immunotherapy-sensitive tumours is merited.

Functional transcription of the *FCU1* transgene, indicative of replicating virus, was evident from pharmacokinetic analyses. Serum 5-FU concentrations ranged from 1 to 1072 ng/ml across all cohorts with significantly elevated levels at higher dose cohorts. Tumour 5-FU titres were detectable in the higher treatment doses, with two patients who received the highest dose of virus having concentrations exceeding 25 pg/mg tissue. The range of 5-FU concentrations from 16 to 35 pg/mg of tissue compared favorably with that of 5.9 ± 0.9 pg/mg reported in tumour tissue of patients with HCC treated with an oral pro-drug of 5-FU [23] and were close to the mean 5-FU concentration of 56.6 pg/mg after *i.t.* injection of a non-propagative vaccinia virus expressing *FCU1* in combination with oral 5-FC [24]. Of interest, *i.v.* administered 5-FU can result in higher serum levels of 5-FU, with targeted serum concentrations of 2,500-3,000 ng/ml [25,26], compared to a median of 82 ng/ml detected in our patients receiving oral 5-FC. Therefore, maximising 5-FU concentrations in the tumour tissue using TG6002/oral 5-FC combination allows direct targeting of malignant cells, whilst minimising systemic toxicity. Higher serum concentrations of 5-FU, as experienced during standard *i.v.* delivery can be problematic, as side effects can be very significant and many patients are unable to tolerate repeated cycles. Moreover, there is frequently rapid development of resistance to 5-FU alone when administered via an *i.v.* route.

Despite the apparent lack of clinical efficacy, early peripheral blood immune cell responses indicated immune activity resulting from the combination therapy; in addition to promoting an anti-tumour response, this may also represent a virally-driven immune response to pave the way for both direct tumour lysis and abscopal effects through immune modulation and 5-FU activation. Calreticulin plasma concentrations increased shortly after TG6002 infusion, peaking 6 hours post-treatment and remaining elevated up to 24 hours. Calreticulin is an endoplasmic reticulum-associated chaperone protein ubiquitously expressed intracellularly, but also released from cells undergoing ICD [27,28] and is one of the main hallmarks of ICD in malignant disease. ICD is a unique class of regulated cell death that elicits antigen-specific adaptive immune processes via release of damage-associated molecular patterns (DAMPs), of which calreticulin is a key component. The overall role of ICD and DAMP release is the recruitment of antigen-presenting cells to the site of dying tumour cells, in order to promote antigen uptake and processing, prior to cross-presentation to T cells to initiate a tumour-specific immune response.

Associated with the occurrence of ICD, mRNA sequencing revealed a significant response of immune cells at the transcriptional level. A considerable elevation in the number of ssDEGs were apparent in patients who received the highest doses of TG6002. These ssDEGs formed clusters representing immune-related pathways in patients receiving higher virus doses at earlier time points than patients receiving lower doses, suggesting greater immune activation at higher doses of virus. GO annotations revealed signaling pathways associated with response to virus involving IFN-stimulated genes (ISGs) and, subsequently, immune activation. Specifically, nine ssDEGs were predominantly upregulated in multiple patients and demonstrated to be highly relevant in the aforementioned pathways.

Immunophenotyping of patient PBMCs evidenced immune cell activation across multiple cell populations. CD69, an early activation marker, was elevated shortly after TG6002 infusion, as was the immune checkpoint ligand PD-L1, a common marker for immune activation and exhaustion following OV therapy [29]. PD-L1 expression following virus therapy is associated with an anti-tumour immune response driven by IFNs and other inflammatory molecules [30]. Furthermore, increased PD-L1 expression by both peripheral and tumour-infiltrating T cells has been associated with better prognosis to immune checkpoint blockade [31,32]. Likewise, PD-1 and TIM-3 expression are indicative of inflammatory cytokine signaling, including inerleukin (IL)-12, IL-15, IL-18 and IFN-γ [33,34]. Elevated expression of OX40 on NK cells is broadly associated with activation, in line with previous data [35]. Engagement of the CD40/CD40L complex is a secondary activation signal required for T cell activation and is associated with enhanced cytokine production by dendritic cells (DCs), coupled with enhanced cross-presentation capacity [36]. This interaction has beneficial effects across the immune system as a whole, and is, therefore, regarded as instrumental in an inflammatory response. Furthermore, the downregulation of CD25 and CTLA-4 on T cells also indicates the immunological switch from suppressive to activation, in response to TG6002/5-FC therapy.

Functional T cell responses to treatment were evident against TG6002 but, more importantly, against TAAs, suggesting an antigen-targeted cytotoxic effect against tumour. TCRβ sequencing revealed that a greater number of T cell clones targeting both cancer antigens in general and specifically neoantigens, emerged within patients receiving higher doses of virus. Anti-cancer clonal T cell expansion is associated with improved anti-cancer activity [37], where increased CD8 T cell tumour infiltration was also observed in the highest dose cohort upon treatment. As such, whether a T cell is activated towards a TAA/neoantigen or the virus itself may ultimately result in a parallel anti-tumour response against virus-infected cells within the tumour microenvironement.

These data, in summary, represent the first in-human dose-escalation study using IHA delivery of a vaccinia virus. This treatment strategy directly targets tumour tissue and is associated with effective viral replication, expression and activity of the *FCU1* transgene, immune activation and evidence for anti-tumour immune activity. Further assessment of IHA delivery of OV in immunotherapy-sensitive cancers is merited.

## Data Availability

All data produced in the present study are available upon reasonable request to the authors

## Declarations

### ClinicalTrials.gov registration

NCT04194034

## Acknowledgments

We are grateful to all the patients that participated in this trial. The research is supported by the National Institute for Health Research (NIHR) infrastructure at Leeds. The views expressed are those of the author(s) and not necessarily those of the NHS, the NIHR or the Department of Health.

Adel Samson acknowledges CRUK grant 29039; consulting fees: Roche, Chugai; academic grants: Replimune, Histosonics, Oncolytic Biotech, Transgene (all paid to institution).

We thank J. Hortelano and M. Gantzer (Transgene) for their assistance with conducting viral plaque assays and RT-qPCR experiments on tumour biopsies. We thank Dr C. Machon and Dr J. Guitton (Service de Biochimie et Pharmacotoxicologie, Hospices Civils de Lyon, France) for quantification of 5-FC, 5-FU and F-BAL on plasma samples and tumour biopsies.

