## Supplementary figures and images for "A phase I clinical trial of intrahepatic artery delivery of TG6002 in combination with oral 5-fluorocytosine in patients with liver-dominant metastatic colorectal cancer"

### Supplementary Figure 1

#### Slide 1
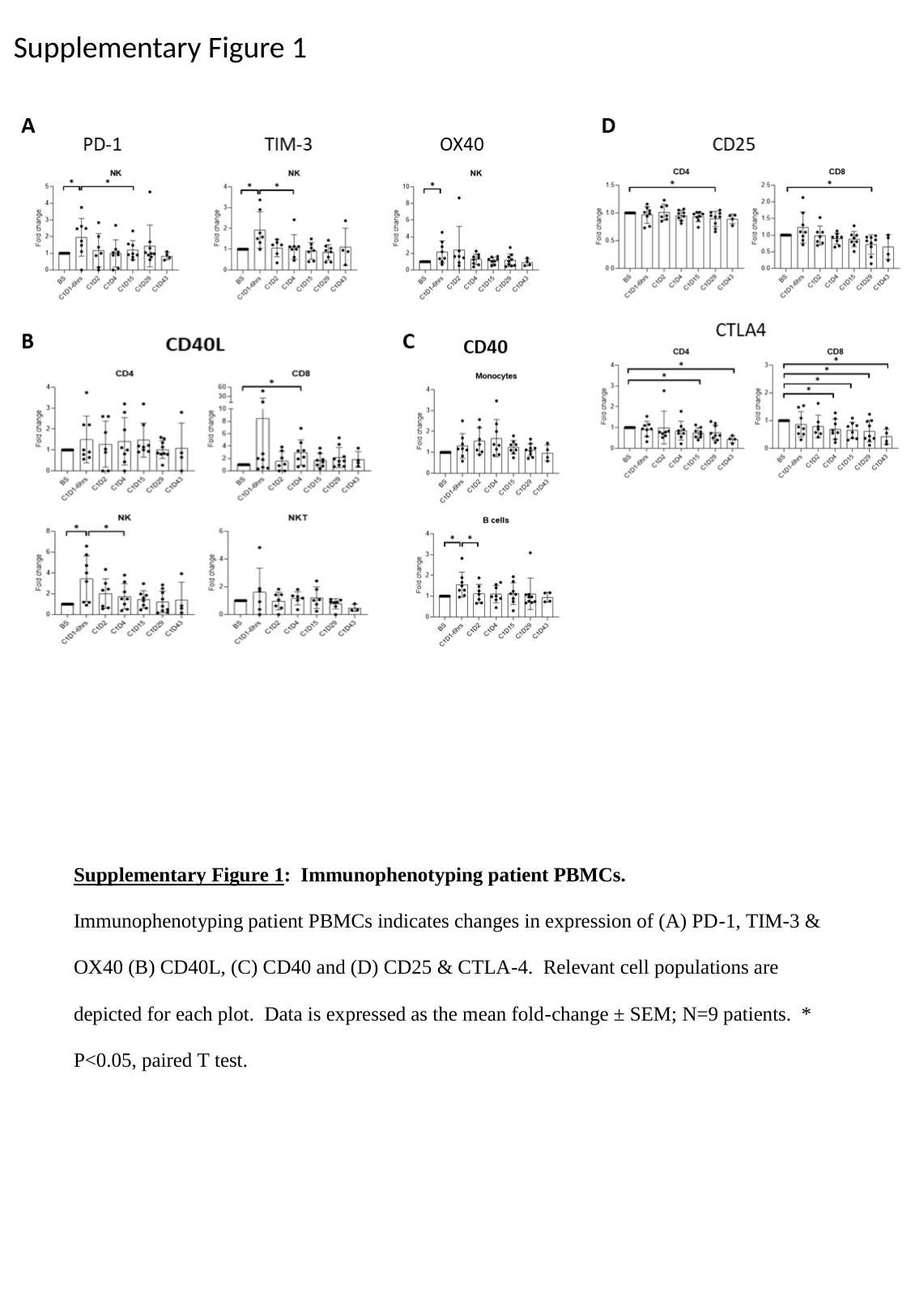

Supplementary Figure 1

### Supplementary Table 2

#### Slide 1
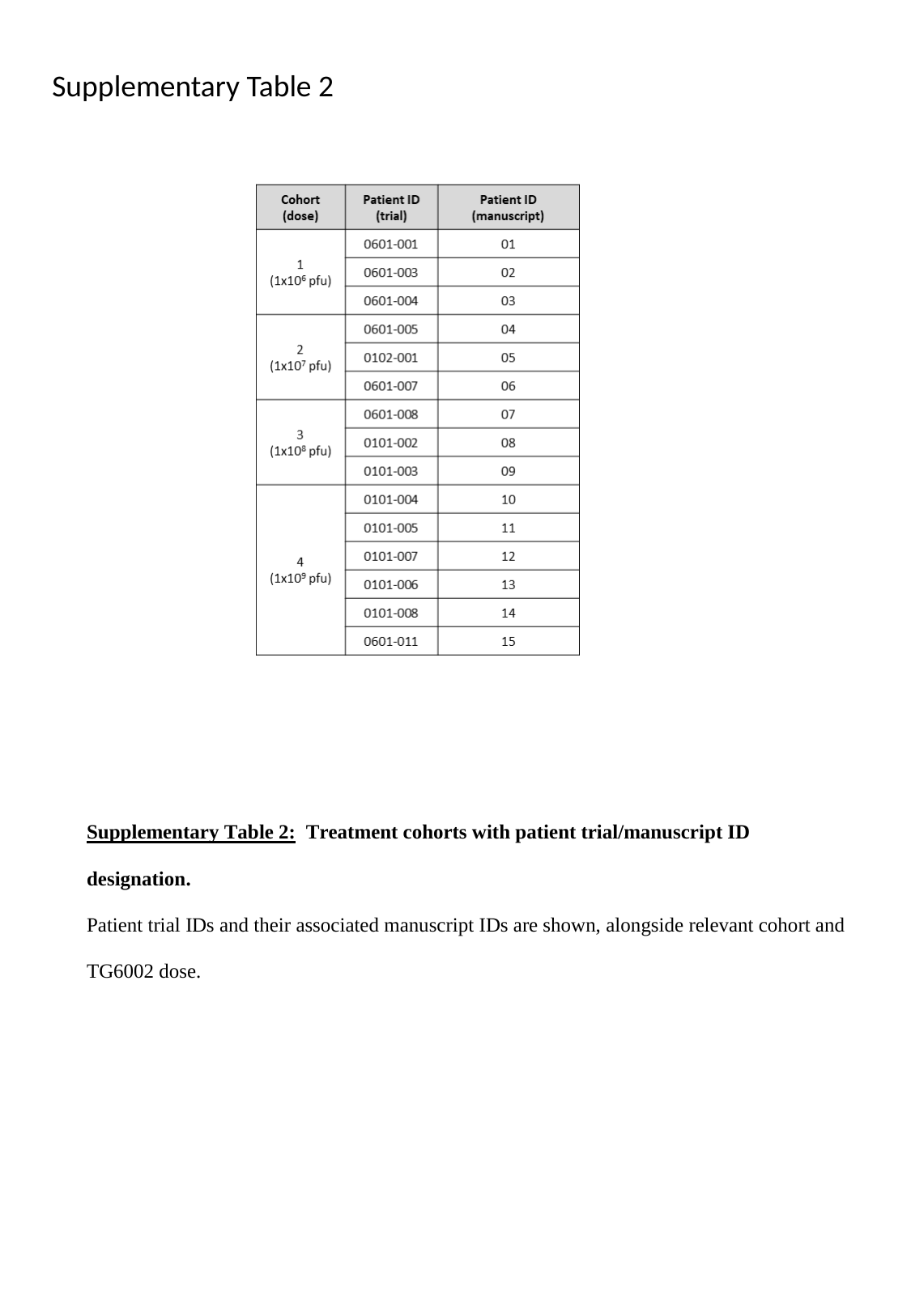

Supplementary Table 2
