## Supplementary Table 1 for "A phase I clinical trial of intrahepatic artery delivery of TG6002 in combination with oral 5-fluorocytosine in patients with liver-dominant metastatic colorectal cancer"

### Slide 1
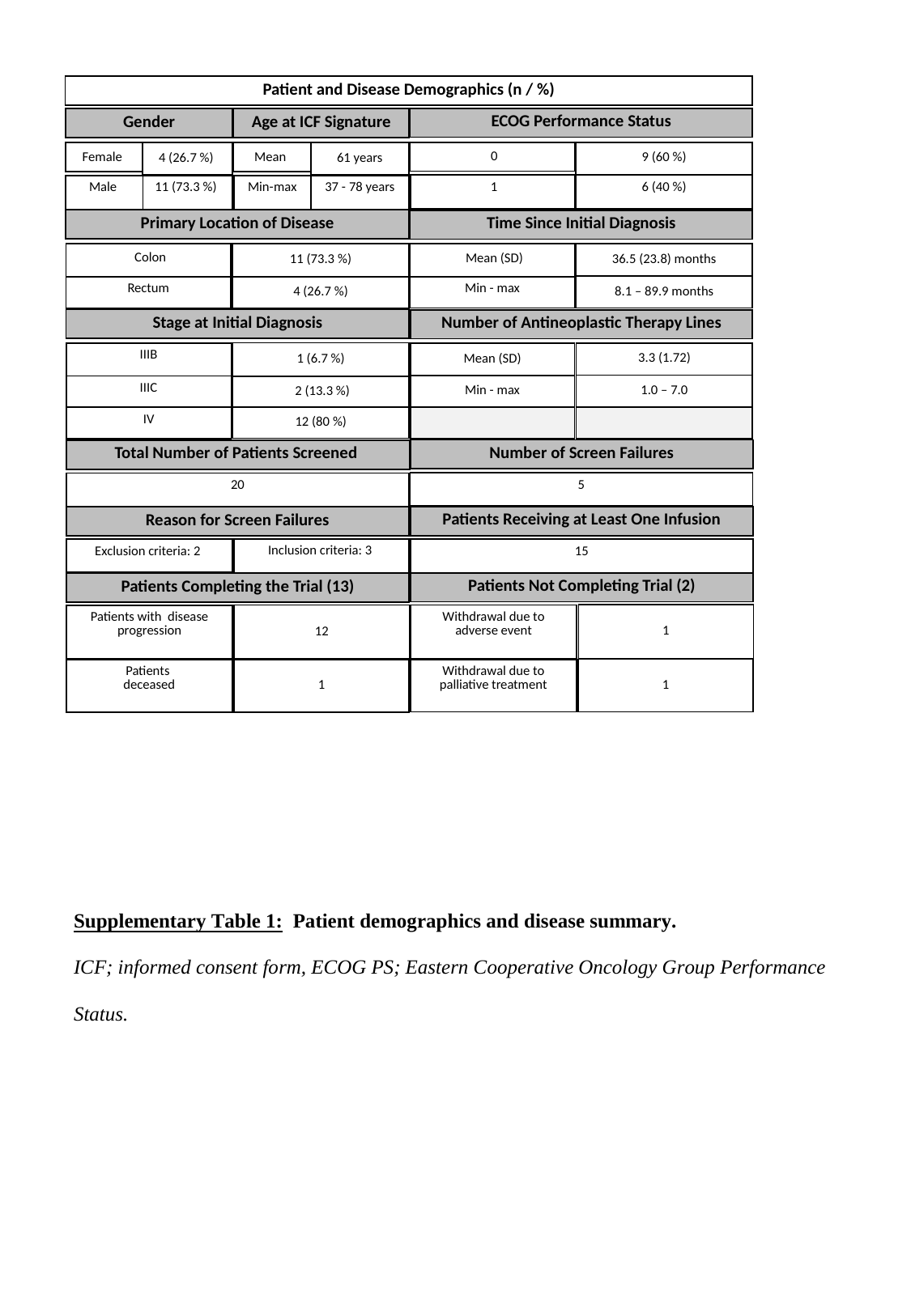

| Patient and Disease Demographics (n / %) |
| --- |
| ECOG Performance Status |
| --- |
| Gender |
| --- |
| Age at ICF Signature |
| --- |
| 0 |
| --- |
| 9 (60 %) |
| --- |
| Female |
| --- |
| 4 (26.7 %) |
| --- |
| Mean |
| --- |
| 61 years |
| --- |
| 1 |
| --- |
| 6 (40 %) |
| --- |
| Male |
| --- |
| 11 (73.3 %) |
| --- |
| Min-max |
| --- |
| 37 - 78 years |
| --- |
| Primary Location of Disease |
| --- |
| Time Since Initial Diagnosis |
| --- |
| Colon |
| --- |
| Rectum |
| 11 (73.3 %) |
| --- |
| 4 (26.7 %) |
| Mean (SD) |
| --- |
| Min - max |
| 36.5 (23.8) months |
| --- |
| 8.1 – 89.9 months |
| Stage at Initial Diagnosis |
| --- |
| Number of Antineoplastic Therapy Lines |
| --- |
| 1 (6.7 %) |
| --- |
| 2 (13.3 %) |
| 12 (80 %) |
| IIIB |
| --- |
| IIIC |
| IV |
| 3.3 (1.72) |
| --- |
| 1.0 – 7.0 |
| Mean (SD) |
| --- |
| Min - max |
| Number of Screen Failures |
| --- |
| Total Number of Patients Screened |
| --- |
| 5 |
| --- |
| 20 |
| --- |
| Patients Receiving at Least One Infusion |
| --- |
| Reason for Screen Failures |
| --- |
| Inclusion criteria: 3 |
| --- |
| 15 |
| --- |
| Exclusion criteria: 2 |
| --- |
| Patients Not Completing Trial (2) |
| --- |
| Patients Completing the Trial (13) |
| --- |
| 1 |
| --- |
| Withdrawal due to adverse event |
| --- |
| 12 |
| --- |
| Patients with disease progression |
| --- |
| 1 |
| --- |
| Withdrawal due to palliative treatment |
| --- |
| 1 |
| --- |
| Patients deceased |
| --- |
